# Course and clinical severity of the SARS-CoV-2 Omicron variant infection in Tianjin, China

**DOI:** 10.1101/2022.06.16.22271932

**Authors:** Yi Ren, Lixia Shi, Yi Xie, Chao Wang, Wenxin Zhang, Feifei Wang, Haibai Sun, Lijun Huang, Yuanrong Wu, Zhiheng Xing, Wenjuan Ren, Joachim Heinrich, Qi Wu, Zhengcun Pei

**Affiliations:** Haihe Hospital, Tianjin University, Tianjin, China; Tianjin Institute of Respiratory Diseases, Tianjin, China; Institute and Clinic for Occupational, Social and Environmental Medicine, University Hospital, Ludwig Maximilians University Munich, Munich, Germany; Allergy and Lung Health Unit, Melbourne School of Population and Global Health, The University of Melbourne, Melbourne, Australia; Medical college, Tianjin University, Tianjin, China

**Keywords:** SARS-Cov-2, Omicron, course, illness severity, children

## Abstract

**Introduction:** There is limited information describing the course and severity of illness in subjects infected by the severe acute respiratory syndrome coronavirus 2 (SARS-CoV-2) Omicron variant, especially in children.

**Methods:** In this population-based cohort study, subjects with Omicron variant infection during the outbreak between January 8 and February 12, 2022 in Tianjin, China were included (n=429). The main outcomes were the distribution of asymptomatic, mild, moderate, and severe patients, and clinical courses including the interval from positive polymerase chain reaction (PCR) test to the onset, aggravation or relief of symptoms, and the interval of reversing positive PCR-test into negative, and length of hospital stay.

**Results:** Of the 429 subjects (113 [26.3%] children; 239 [55.7%] female; median age, 36 years [IQR 15.0 to 55.0 years]), the proportion (95% CI) of symptomatic subjects on admission was 95.6% (93.2%, 97.2%), including 60.4% (55.7%, 64.9%) mild, 35.0% (30.6%, 39.6%) moderate, and 0.2% (0.0%, 1.3%) severe. Compared with adults, children had lower proportion of moderate Covid-19 (8.8% vs 44.3%). On discharge, 45.9% (41.3%, 50.7%) and 42.2% (37.6%, 46.9%) of the subjects were diagnosed as having experienced mild and moderate Covid-19. The median (IQR) length of hospital stay was 14.0 (12.0, 15.0) days. The median interval of reversing positive PCR-test into negative was 12.0 (10.0, 13.0) days.

**Discussion:** Symptomatic and moderate Covid-19 in Omicron infections was common in adults and children, recovery from Omicron infections took around 2 weeks of time. The SARS-CoV-2 Omicron infection in this study was not as mild as previously suggested.

**Key Messages:** *What is already known on this topic:* Previous studies have demonstrated that Omicron patients were more likely to be mild, less hospitalized, less severe disease, and of lower mortality. However, there is limited information describing the course and severity of illness in subjects infected by the Omicron variant, especially in children.

*What this study adds:* This study reveals that the prevalence of symptomatic and moderate Covid-19 in Omicron infections was considerably high for children and adults in China. In this population based cohort study of 113 children and 316 adults with Omicron variant infection contracted during the outbreak in Tianjin, China, 95.6% of the subjects were symptomatic on admission. Although children had significantly lower proportions of moderate Covid-19 on admission compared to adults (8.8% vs 44.3%), almost one of ten infected children suffered from moderate COVID-19.

*How this study might affect research, practice or policy:* This study expands our understanding of the course and illness severity of the SARS-CoV-2 Omicron infections, especially in children. Awareness and appropriate control policies are needed to reduce moderate illnesses by the Omicron infections.

## Introduction

Omicron, the fifth severe acute respiratory syndrome coronavirus 2 (SARS-CoV-2) variant of concern (VoC) [1], has become the dominant variant in many countries since first sequenced in November 2021. On January 8, 2022, the first laboratory-confirmed Omicron case in mainland China emerged in Tianjin after a successful null COVID strategy in China. Soon after, over 400 Omicron-infected patients were identified through 4 rounds of whole-population screening of approximately 14 million inhabitants. As of February 12, 2022, nearly 90% of the cases had recovered and discharged from the designated hospital.

Limited information has been available to describe the severity and the course of illness in patients infected by the Omicron variant, especially in children. Reduced illness severity of Omicron infection has been reported in South Africa, England and Canada [2-5], however, the policy of Covid-19 control and prevention, population demographics, extend of previous infection, vaccination characteristics are substantially different in China.

This study describes the illness severity and course of SARS-CoV-2 Omicron infection in a large general population of children and adults residing in the metropolitan area of Tianjin, using data of all identified infected subjects during the outbreak.

## Materials and methods

### Study design and population

This population-based cohort study was conducted in Tianjin, where the first Omicron outbreak in China took place. The metropolitan area of Tianjin has a population size of 14 million. Since the identification of the first patient with Omicron infection on January 8, 2022, Tianjin instituted a fast and comprehensive testing policy, where 4 rounds of whole-population screening were conducted to identify persons with positive result on polymerase chain reaction (PCR) testing of a nasopharyngeal or pharyngeal swab sample. As of January 21, 2022, the total number of PCR tests exceeded 58 million and all inhabitants were tested at least once (Figure 1). The positive tested subjects were transferred to and monitored at Haihe Hospital, the only designated hospital for Covid-19 patients in Tianjin. Persons who had close contact with the positive tested subjects for 2 days before the occurrence of symptoms, or 2 days before the sampling of asymptomatic infected subjects, were transferred to designated hotels and started a 14-day quarantine thereafter. They were retested every day and were transferred to hospital if they got a positive test result, or sent back home if they remained test negative by the end of quarantine. All Omicron infections were finally confirmed by Chinese center for disease control and prevention with whole genome sequencing. In total, 429 consecutive, all positive tested subjects admitted between January 8 and February 12, 2022, including children and adults, were included in this analysis.

**Figure 1.**
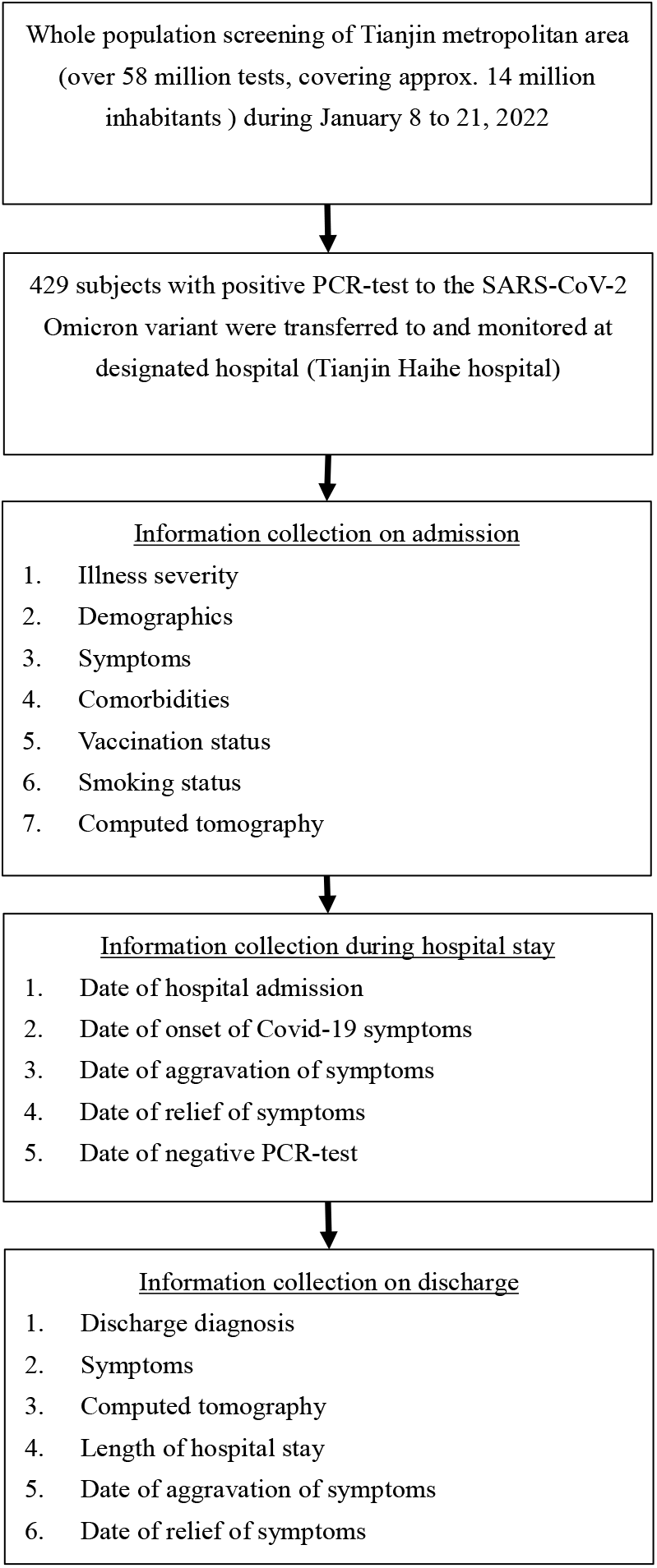
Flow chart of patient screening and information collection.

Subjects below 18 years were labeled as “children” in the present texts. This study was approved by the institutional review board at Haihe Hospital (Approval number: 2022HHWZ-002), the written informed consent was waived owing to the use of retrospective data.

### Study outcomes and data collection

The primary outcome of interest was illness severity and its progression during hospital stay, including asymptomatic, mild, moderate, and severe patients at admission to hospital and at discharge. Several clinical course-related time intervals were used as secondary outcomes, including the interval from hospital admission to the onset of Covid-19 sympt<u>o</u>ms in presymptomatic patients; the interval from hospital admission to aggravation (including development of new symptoms or worsen of preexisting symptoms) or relief of symptoms for the first time in symptomatic patients, and the interval of reversing positive PCR-test into negative, and length of hospital stay of all patients.

Illness severity was defined by clinical physicians based on the latest version of Chinese guidelines for the diagnosis and treatment of Covid-19 [6], which is highly consistent with the World Health Organization criteria [7]. Briefly, mild disease was defined as presenting mild symptoms, including fever, cough, sore throat, sputum, runny nose, myalgias, etc., but without evidence of viral pneumonia or hypoxia. Moderate disease was defined as having non-severe signs of pneumonia, such as fever, cough, dyspnea, fast breathing, chest indrawing, etc. In addition, moderate disease was distinguished from mild disease assisted with chest Computed Tomography (CT) by confirming the appearance of pneumonia, such as ground-glass opacity, nodular lesion, consolidation, infiltration, etc. Presymptomatic was defined as being asymptomatic on admission but developed symptoms during hospital stay. Vaccination status was defined as unvaccinated, partially vaccinated (one dose), fully vaccinated (two doses) or booster vaccinated (two priming doses plus a booster dose). The inhabitants of Tianjin received exclusively inactivated vaccines, namely BBIBP-CorV [8] or CoronaVac [9,10], which were manufactured in China. Both vaccines were approved as highly effective against symptomatic COVID-19, COVID-19 pneumonia, and severe COVID-19 [11,12]. Information on demographic characteristics, comorbidities (i.e. cardiovascular disease, diabetes, endocrine disorder, respiratory disease, and tumor), clinical features, radiographical appearances were extracted from patients’ electronic medical record, and the intervals from admission to specific events were manually calculated by via medical record review.

### Statistical analysis

Analyses were stratified by age (children: < 18 years vs adults: ≥ 18 years), or by illness severity status on admission and discharge diagnosis (moderate vs mild) to compare the differences between groups. Asymptomatic and severe diseases were excluded from some analyses because of small numbers in subgroups.

Continuous variables included in the present analyses were not normally distributed, thus data were described with median (interquartile range, IQR), and categorical variables with n (%). The distribution of illness severity was described with proportions (95% confidence interval, 95%CI). Continuous variables were compared between groups using Mann-Whitney U test or Kruskal-Wallis test, as appropriate. Categorical variables were compared using a χ² test or Fisher’s exact test, as appropriate. Multivariable logistic and linear regression models were used to calculate crude and adjusted odds ratios (OR) or β-coefficients and their 95% CIs for the associations between illness severity, or course intervals and their influencing factors, including age group (<18 years vs ≥ 18 years), sex, BMI, vaccination status, smoking, and comorbidities of cardiovascular diseases and diabetes, etc. Illness severity was additionally adjusted for the course interval models. The categories of unvaccinated and partially vaccinated were combined as reference because of small numbers. A 5% significance level (2-sided) was used. Analysis was performed using version 4.1.2 of the R programming language (R Project for Statistical Computing; R Foundation).

## Results

### Clinical characteristics of study population

A total of 429 patients were included (median age 36.0 years; IQR 15.0 to 55.0; range 0 to 90 years; 55.7% female), 113 (26.3%) were children (median age 10.0 years; IQR 8.0 to 11.0; 51.3% female) and 316 (73.7%) were adults (median age 47.0 years; IQR 34.0 to 58.0; 51.3% female) (Table 1). Eighty out of 113 (70.8%) children had received two doses of vaccines and only one child had received a booster injection, while 106 (33.5%) and 142 (44.9%) adults had received two or three doses. Children had fewer comorbidities (2.7% vs 32.0%, p<0.001) and fewer abnormal lung findings in chest CT on admission (47.8% vs 84.8%, p<0.001) as compared with adults. As of the present analyses, there were still 44 (10.3%) patients hospitalized (3 children and 41 adults).

**Table 1.** Demographic and radiographical findings of subjects with Omicron infection, by age.

|  | Overall (n=429) | Children (n=113) | Adults (n=316) | p-value |
| --- | --- | --- | --- | --- |
| Age, years | 36.0 (15.0, 55.0) | 10.0 (8.0, 11.0) | 47.0 (34.0, 58.0) | <0.001 |
| Sex |  |  |  | 0.274 |
| Male | 190 (44.3) | 55 (48.7) | 135 (42.7) |  |
| Female | 239 (55.7) | 58 (51.3) | 181 (57.3) |  |
| Weight, kg | 65.0 (52.0, 77.0) | 36.0 (25.50, 51) | 70.0 (60, 80) | <0.001 |
| Height, cm | 162.0 (155.0, 170.0) | 145.0 (130.5, 157.5) | 167.0 (160.0, 172.0) | <0.001 |
| BMI, kg/m<sup>2</sup> | 24.0 (20.5, 27.3) | 18.0 (15.4, 22.043) | 25.4 (22.4, 28.1) | <0.001 |
| Vaccination status |  |  |  | 0.009 |
| Unvaccinated | 22 (5.1) | 9 (8.0) | 13 (4.1) | 0.135 |
| Partially vaccinated | 23 (5.4) | 6 (5.3) | 17 (5.4) | 1.000 |
| Fully vaccinated | 186 (43.4) | 80 (70.8) | 106 (33.5) | <0.001 |
| Booster vaccinated | 143 (33.3) | 1 (0.9) | 142 (44.9) | <0.001 |
| Unknown | 55 (12.8) | 17 (15.0) | 38 (12.0) | 0.415 |
| Current or former smoker | 38 (8.9) | 0 (0.0) | 38 (12.0) | - |
| Comorbidities |  |  |  |  |
| Any | 104 (24.2) | 3 (2.7) | 101 (32.0) | <0.001 |
| Cardiovascular disease | 89 (20.7) | 2 (1.8) | 87 (27.5) | <0.001 |
| Endocrine disorder | 5 (1.2) | 0 (0.0) | 5 (1.6) | - |
| Diabetes | 31 (7.2) | 1 (0.9) | 30 (9.5) | 0.001 |
| Respiratory disease | 7 (1.6) | 0 (0.0) | 7 (2.2) | - |
| Tumor | 2 (0.5) | 0 (0.0) | 2 (0.6) | - |
| Computed Tomography findings on admission |  |  |  |  |
| Any abnormal lung findings | 322 (75.1) | 54 (47.8) | 268 (84.8) | <0.001 |
| Lateral | 121 (28.2) | 27 (23.9) | 94 (29.7) | 0.240 |
| Bilateral | 190 (44.3) | 20 (17.7) | 170 (53.8) | <0.001 |
| Ground-glass opacities | 144 (33.6) | 13 (11.5) | 131 (41.5) | <0.001 |
| Consolidation | 6 (1.4) | 0 (0.0) | 6 (1.9) | - |
| Nodular lesion | 193 (45.0) | 32 (28.3) | 161 (50.9) | <0.001 |
| Infiltration | 67 (15.6) | 10 (8.8) | 57 (18.0) | 0.031 |

<sup>a</sup> Asymptomatic at discharge denotes being asymptomatic during the entire hospital stay
<sup>b</sup> Symptomatic at discharge included patients who were symptomatic or presymptomatic on admission
Data are median (IQR) or n (%). p values were calculated by Mann-Whitney U test, Kruskal-Wallis test, $\chi^2$ test, or Fisher's exact test, as appropriate. $\chi^2$ test comparing all subcategories.

The most common symptoms in children and adults were fever and cough, followed with sputum and sore throat without differences (Table 2 in the Supplement). Half of children with moderate Covid-19 had runny nose, which was higher than that for adults (50.0% vs 5.7%, p < 0.001). Within the mild Covid-19 group, the proportion of abnormal CT findings on admission in children were lower than that in adults, with the exception of infiltration; while within the moderate group, there were no major differences between the two groups. On discharge, children who experienced moderate Covid-19 had higher proportion of lateral abnormal CT findings than that for adults (50.0% vs 12.1%, p = 0.007).

**Table 2.**
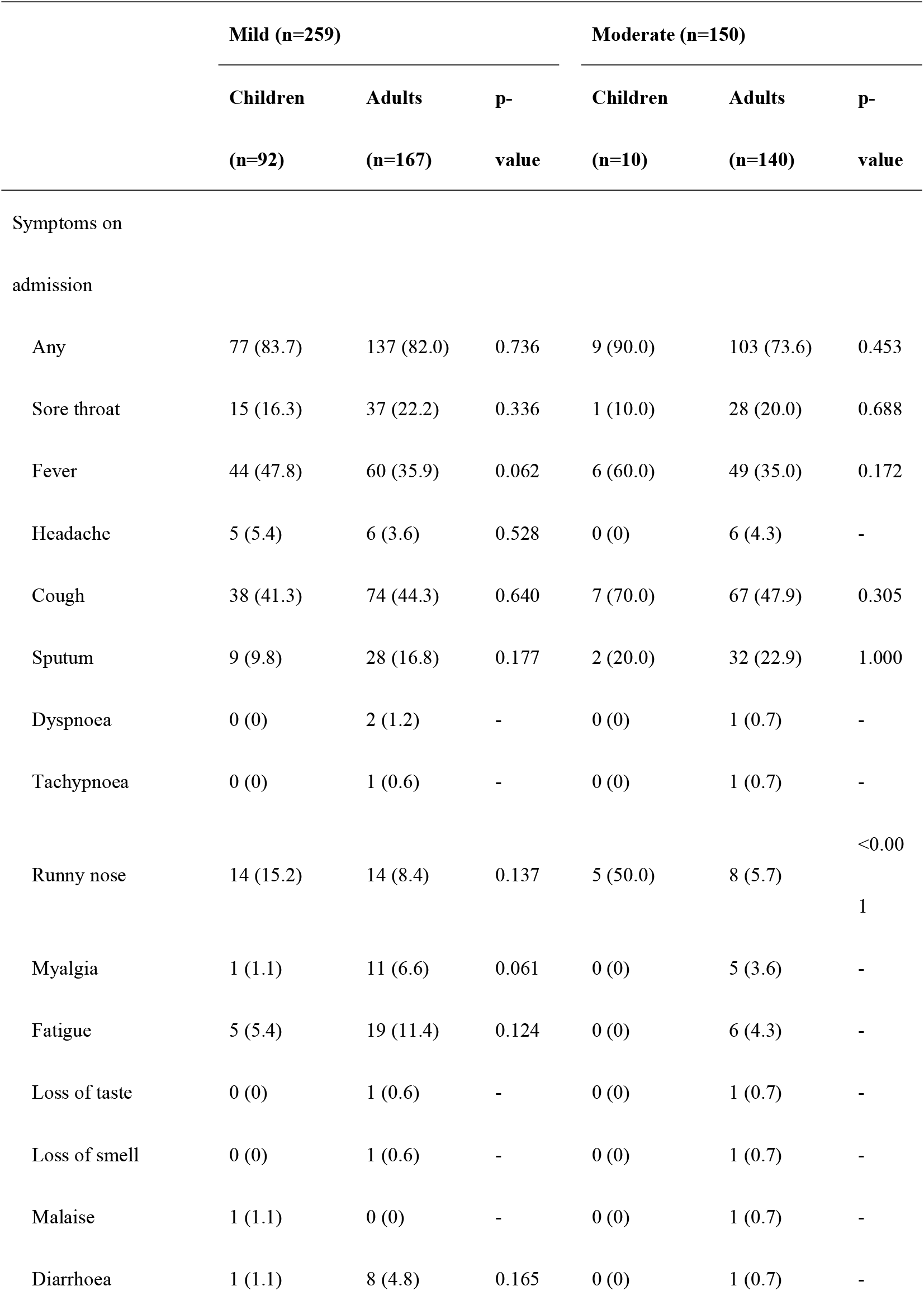
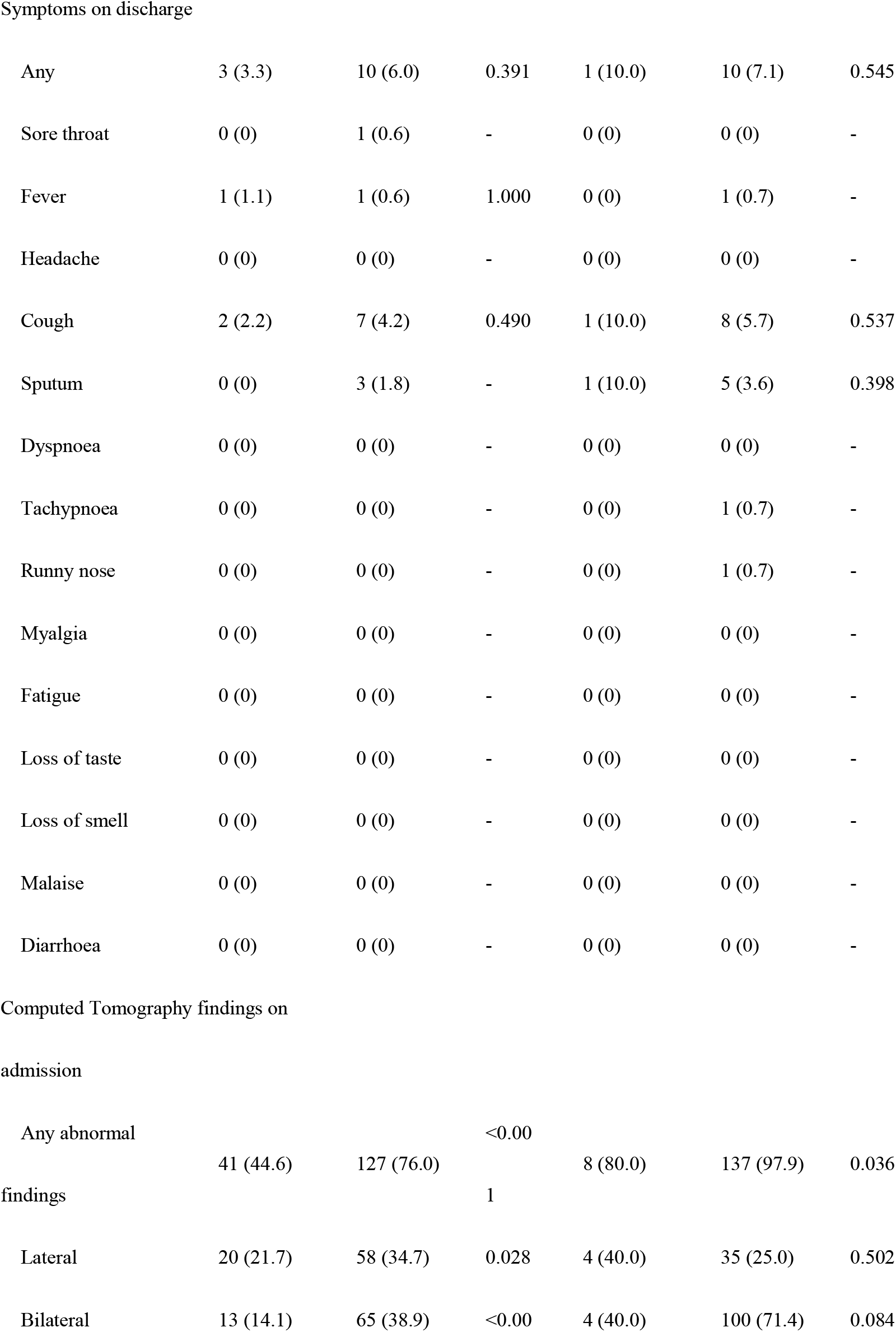
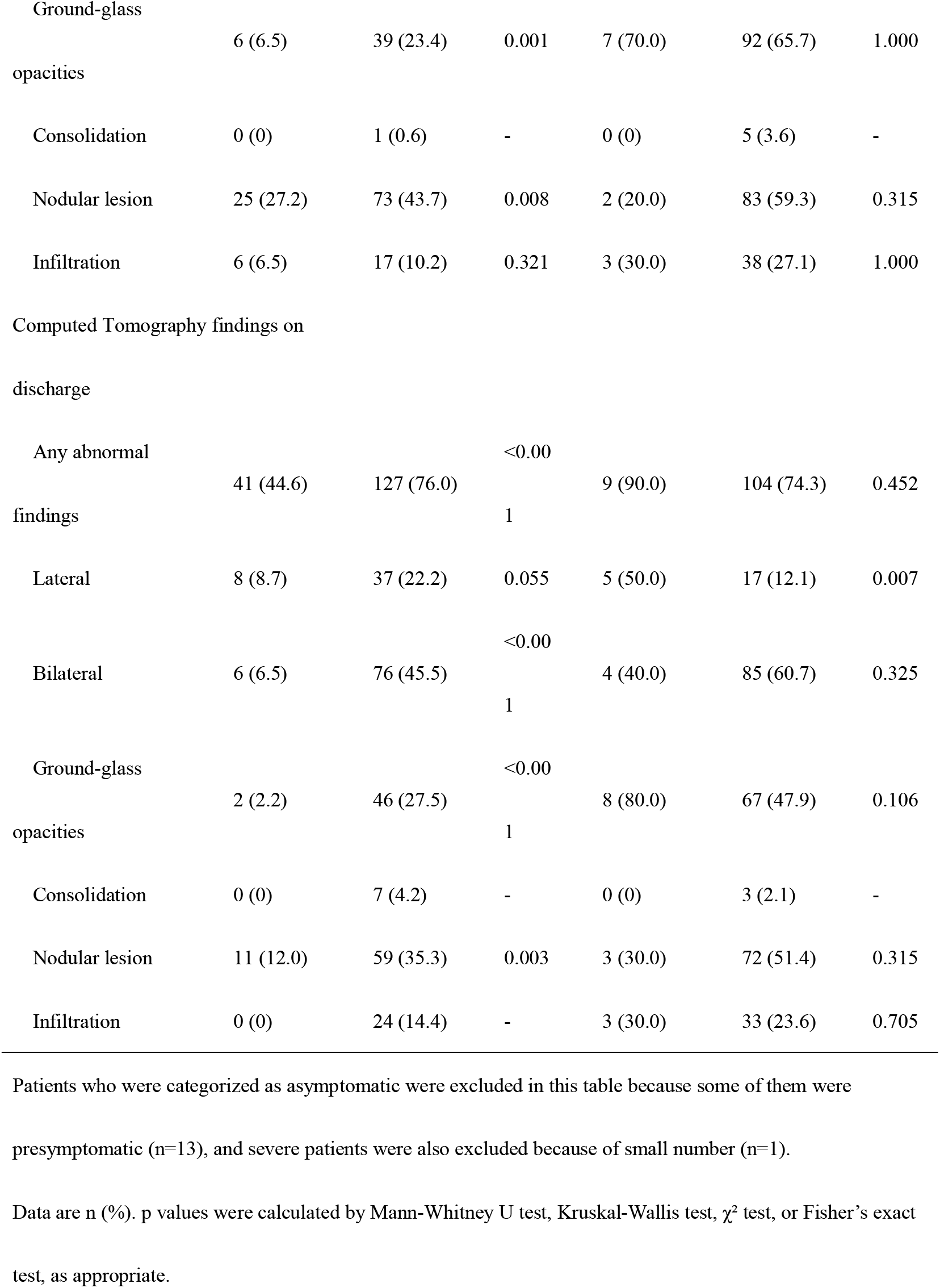
Clinical and radiographical findings in mild and moderate Omicron infections, by age.

### Illness severity of the SARS-CoV-2 Omicron variant

On admission, 19 out of 429 subjects (4.4% [95%CI: 2.9%, 6.8%]) were asymptomatic and 95.6% (93.2%, 97.2%) were symptomatic and developed at least a mild course, including 60.4% (55.7%, 64.9%) mild, 35.0% (30.6%, 39.6%) moderate, and 0.2% (0.0%, 1.3%) severe (Table 3), whereas 45.9% (41.3%, 50.7%) of the subjects were diagnosed as having experienced mild Covid-19 on discharge, 42.2% (37.6%, 46.9%) experienced moderate Covid-19. During the hospital stay, 6 (1.4% [0.6%, 3.0%]) subjects remained asymptomatic. Compared with adults, children had higher proportions of asymptomatic (9.7% vs 2.5%, p = 0.003) and mild Covid-19 (81.4% vs 52.8%, p < 0.001), and lower proportion of moderate Covid-19 (8.8% vs 44.3%, p < 0.001) on admission. On discharge, children had milder illness severities as compared with adults.

**Table 3.**
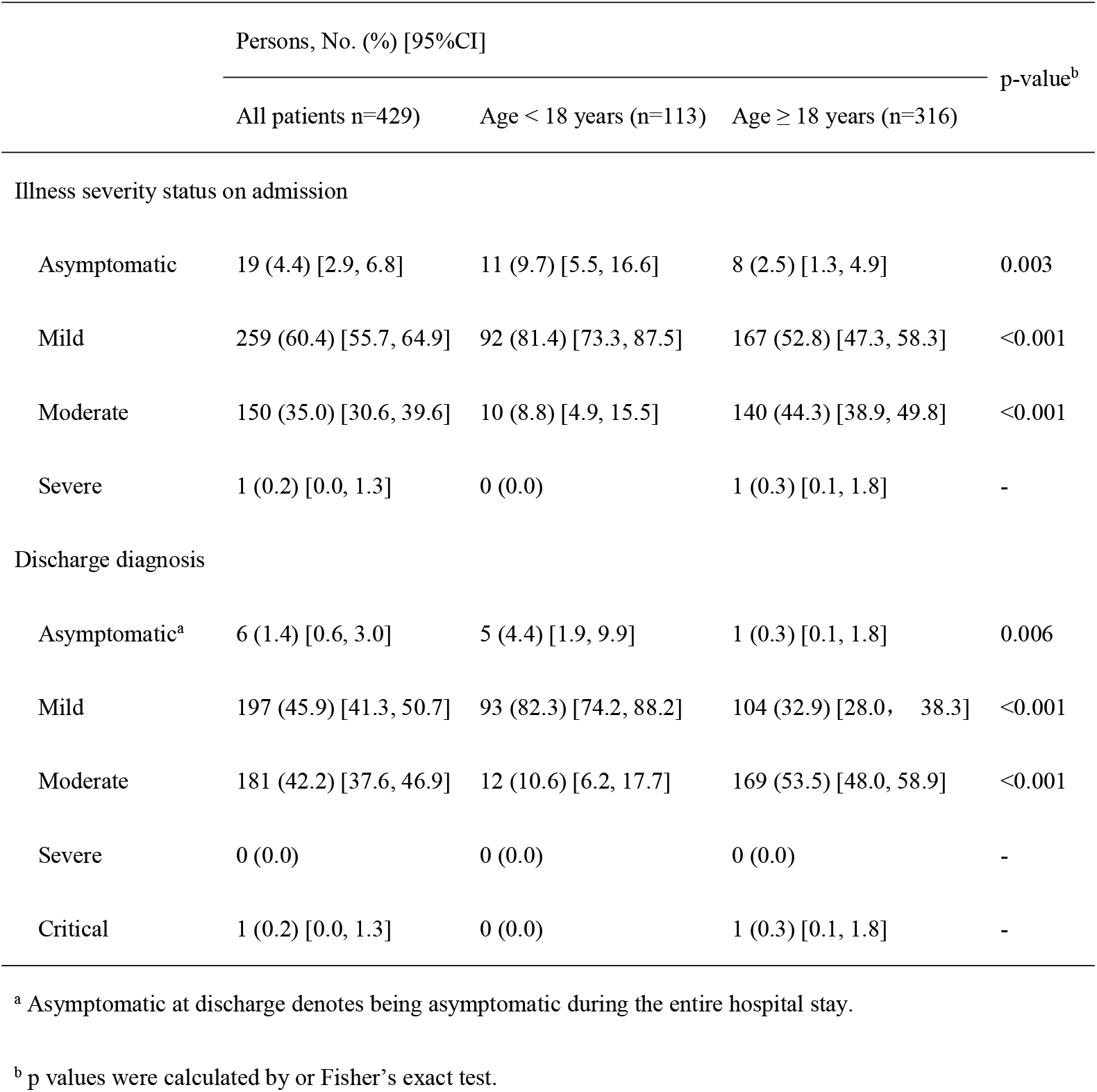
Distribution of illness severity of persons infected with the SARS-CoV-2 Omicron variant.

|  | Persons, No. (%) [95%CI] |  |  | p-value<sup>b</sup> |
| --- | --- | --- | --- | --- |
|  | All patients n=429) | Age < 18 years (n=113) | Age ≥ 18 years (n=316) |  |
| Illness severity status on admission |  |  |  |  |
| Asymptomatic | 19 (4.4) [2.9, 6.8] | 11 (9.7) [5.5, 16.6] | 8 (2.5) [1.3, 4.9] | 0.003 |
| Mild | 259 (60.4) [55.7, 64.9] | 92 (81.4) [73.3, 87.5] | 167 (52.8) [47.3, 58.3] | <0.001 |
| Moderate | 150 (35.0) [30.6, 39.6] | 10 (8.8) [4.9, 15.5] | 140 (44.3) [38.9, 49.8] | <0.001 |
| Severe | 1 (0.2) [0.0, 1.3] | 0 (0.0) | 1 (0.3) [0.1, 1.8] | - |
| Discharge diagnosis |  |  |  |  |
| Asymptomatic<sup>a</sup> | 6 (1.4) [0.6, 3.0] | 5 (4.4) [1.9, 9.9] | 1 (0.3) [0.1, 1.8] | 0.006 |
| Mild | 197 (45.9) [41.3, 50.7] | 93 (82.3) [74.2, 88.2] | 104 (32.9) [28.0, 38.3] | <0.001 |
| Moderate | 181 (42.2) [37.6, 46.9] | 12 (10.6) [6.2, 17.7] | 169 (53.5) [48.0, 58.9] | <0.001 |
| Severe | 0 (0.0) | 0 (0.0) | 0 (0.0) | - |
| Critical | 1 (0.2) [0.0, 1.3] | 0 (0.0) | 1 (0.3) [0.1, 1.8] | - |

<sup>a</sup> Asymptomatic at discharge denotes being asymptomatic during the entire hospital stay.
<sup>b</sup> p values were calculated by or Fisher's exact test.

Compared with children, adults were more likely to have moderate Covid-19 than mild on admission (odds ratio [OR]: 4.95, 95%CI: 2.20, 11.97) (Table 4), and were more likely to experience moderate than mild during hospital stay (OR: 7.95, 95%CI: 3.68, 18.23). In adults, the OR for age and illness severity (moderate vs mild) was 1.04 (95% CI: 1.02,1.06) on admission and 1.05 (95% CI: 1.02, 1.07) on discharge. BMI was positively associated with illness severity in discharge diagnosis (OR, 1.08, 95%CI: 1.01, 1.16).

**Table 4.**
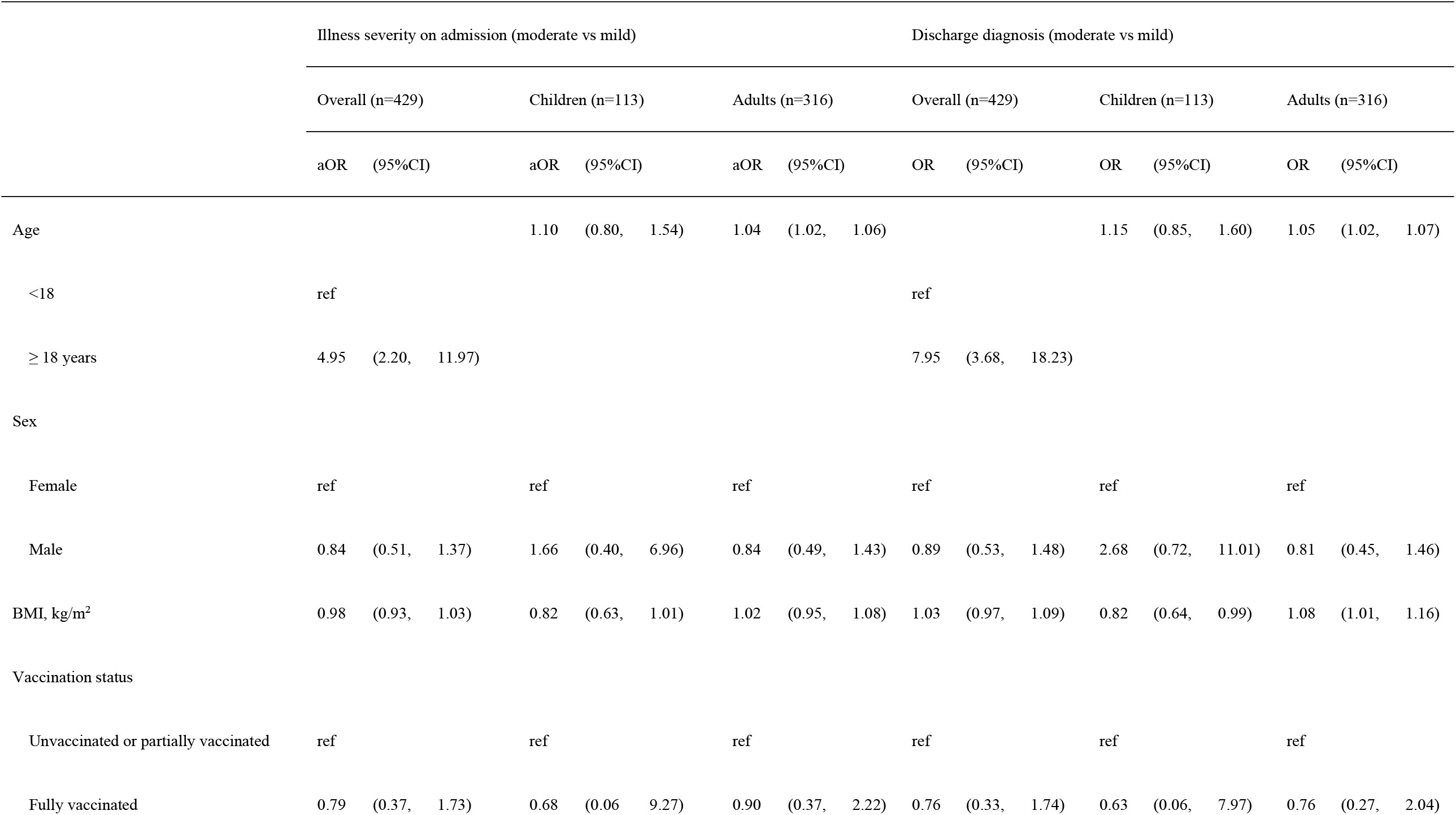
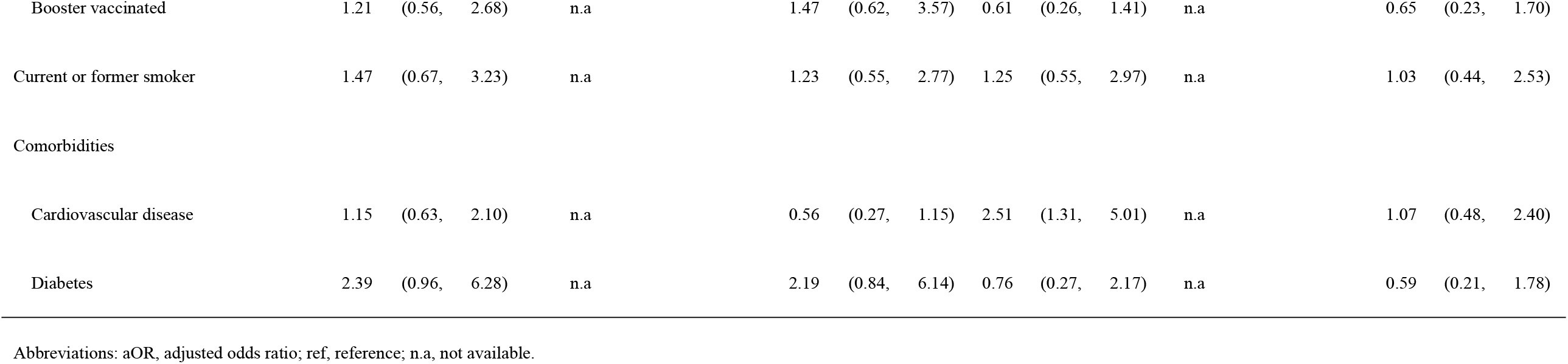
Multivariable logistic regression with illness severity or discharge diagnosis as dependent variable stratified by age.

### Course of the SARS-CoV-2 Omicron Variant

The median length of hospital stay was 14.0 (IQR: 12.0, 15.0) days for all patients, there was no difference between children and adults (median: 13.5, IQR: 12.0, 15.0 and median: 14.0, IQR: 12.0, 16.0, p = 0.21) (Figure 2). The median interval of reversing positive PCR-test into negative was 12.0 (10.0, 13.0) days. The median interval from hospital admission to the onset of Covid-19-relevant symptoms in presymptomatic patients was 4.0 (2.0, 5.0) days; the median intervals from hospital admission to aggravation of symptoms for patients with mild and moderate were 3.0 (2.0, 5.0) days and 4.0 (3.0, 5.0) days, respectively. The median intervals from hospital admission to relief of symptoms for presymptomatic, mild and moderate severity were 9.0 (8.0, 10.0), 5.0 (3.0, 6.0) and 5.0 (4.0, 7.0) days, respectively. Compared with unvaccinated or partially vaccinated patients, adults who received three doses of vaccines had shorter interval of reversing positive PCR-test into negative (β: -1.59, 95%CI: -2.99, -0.20) (Table 5). Children with moderate illness on admission were more likely to have delayed aggravation of Covid-19 symptoms (β: 2.72, 95%CI: 0.11, 5.32). In adults, BMI and cardiovascular disease were associated with higher odds of aggravation of Covid-19 symptoms (OR: 1.06, 95%CI: 1.00, 1.13; and OR: 2.18, 95%CI: 1.07, 4.52, respectively) (Table 6 in the Supplement).

**Table 5.**
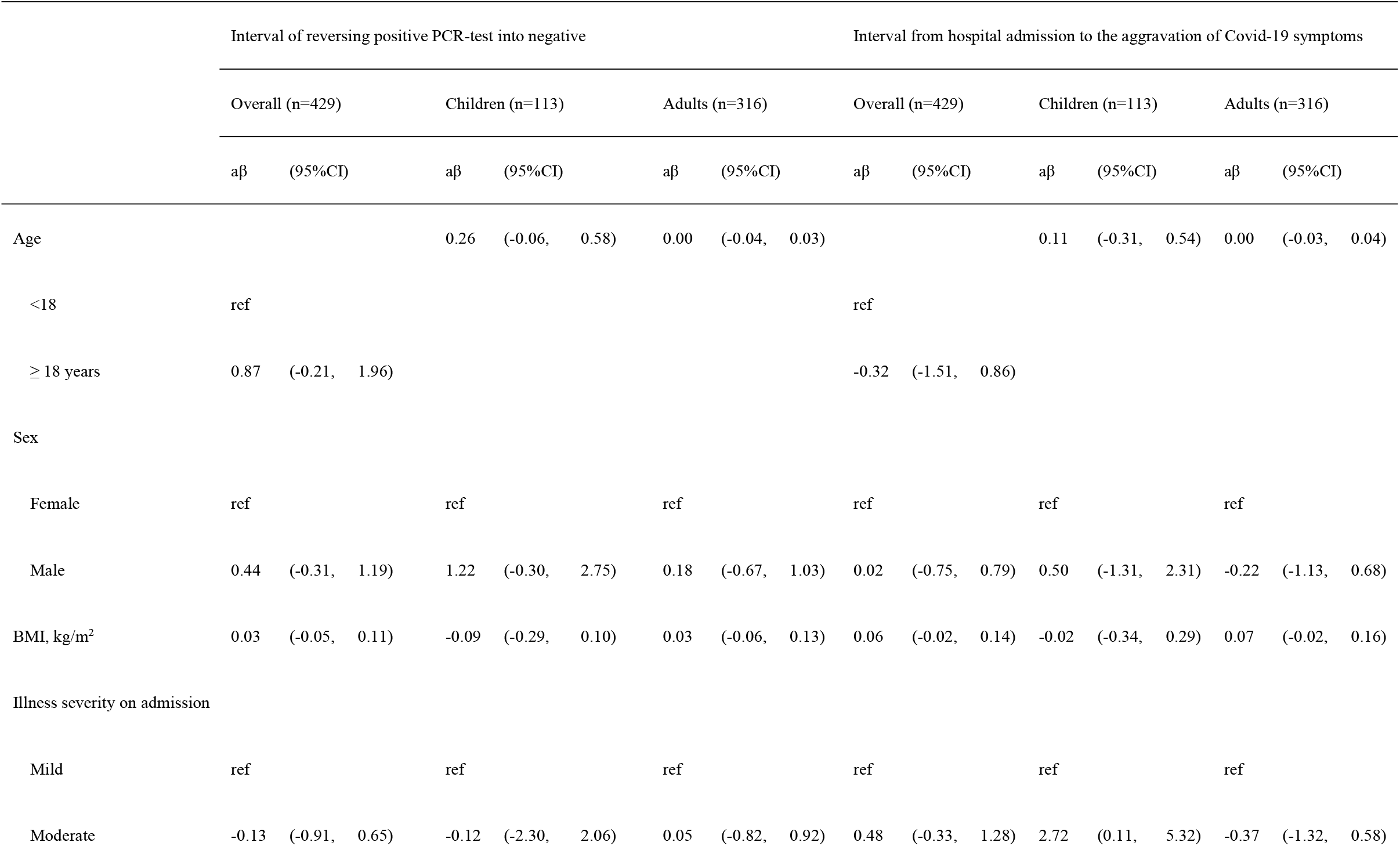
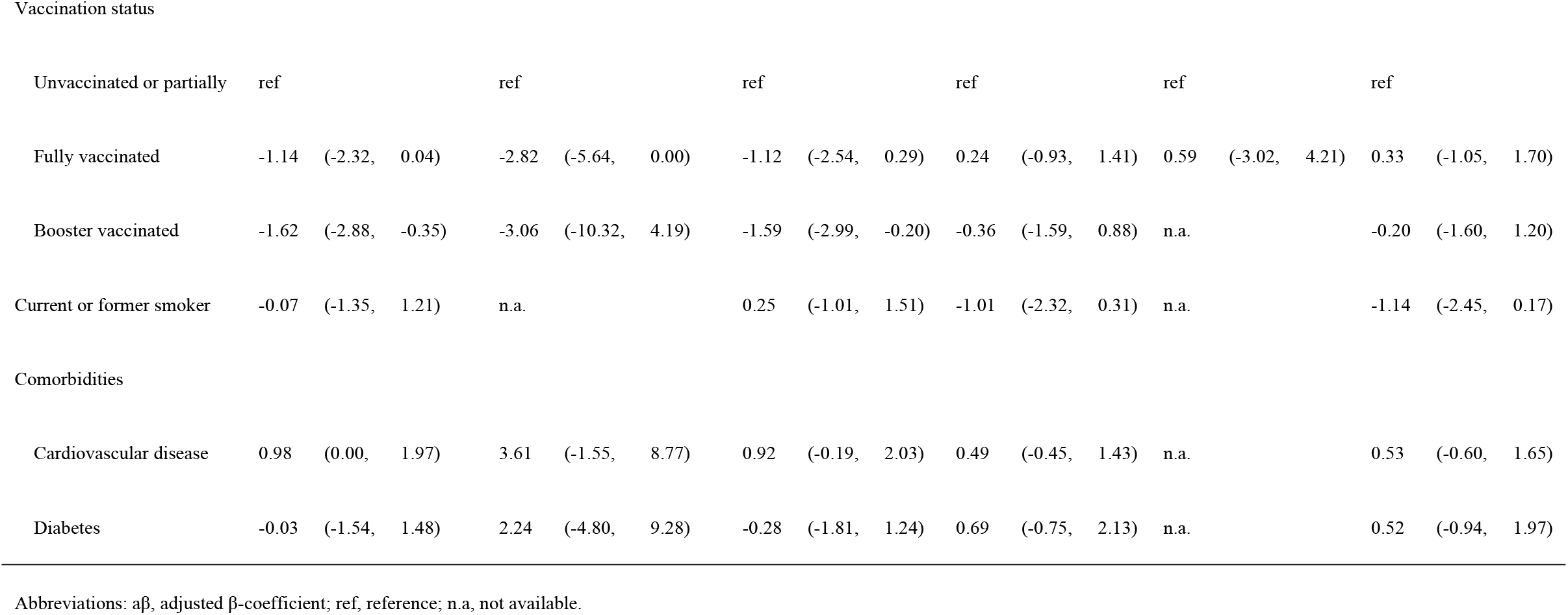
Multivariable linear regression with course-related intervals of the Omicron variant as dependent variable stratified by age.

**Table 6.**
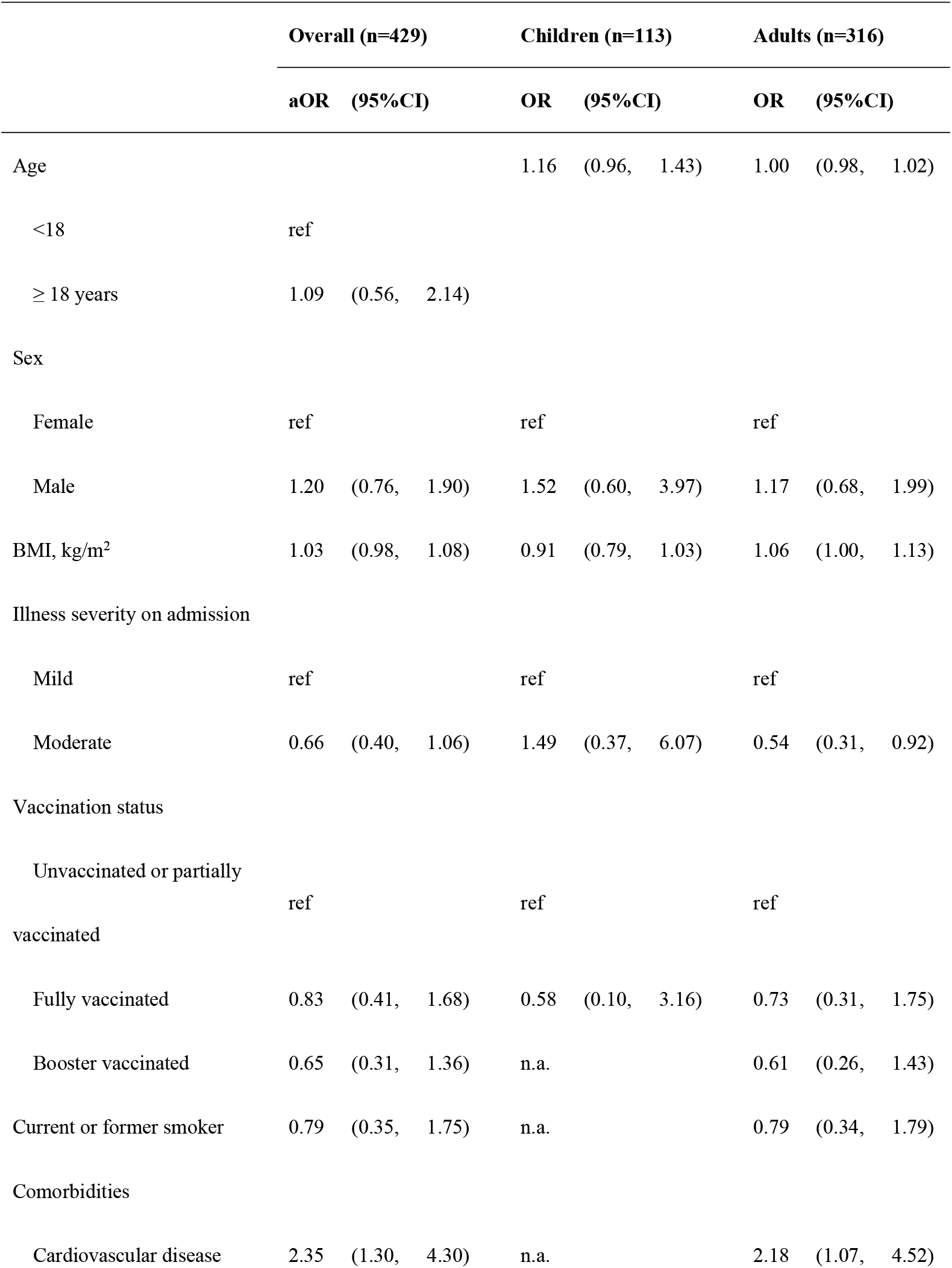
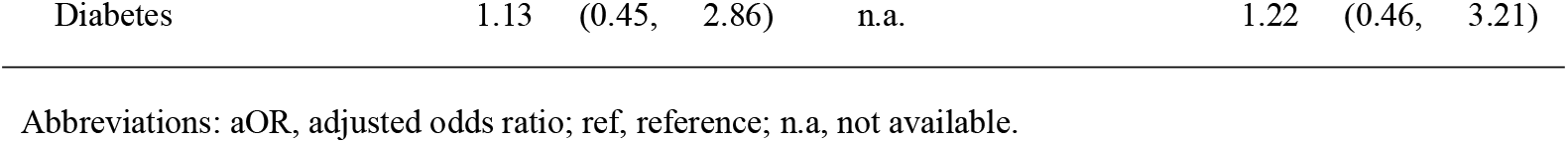
Multivariable logistic regression with aggravation of symptoms of the Omicron variant during hospital stay as dependent variable stratified by age.

**Figure 2.**
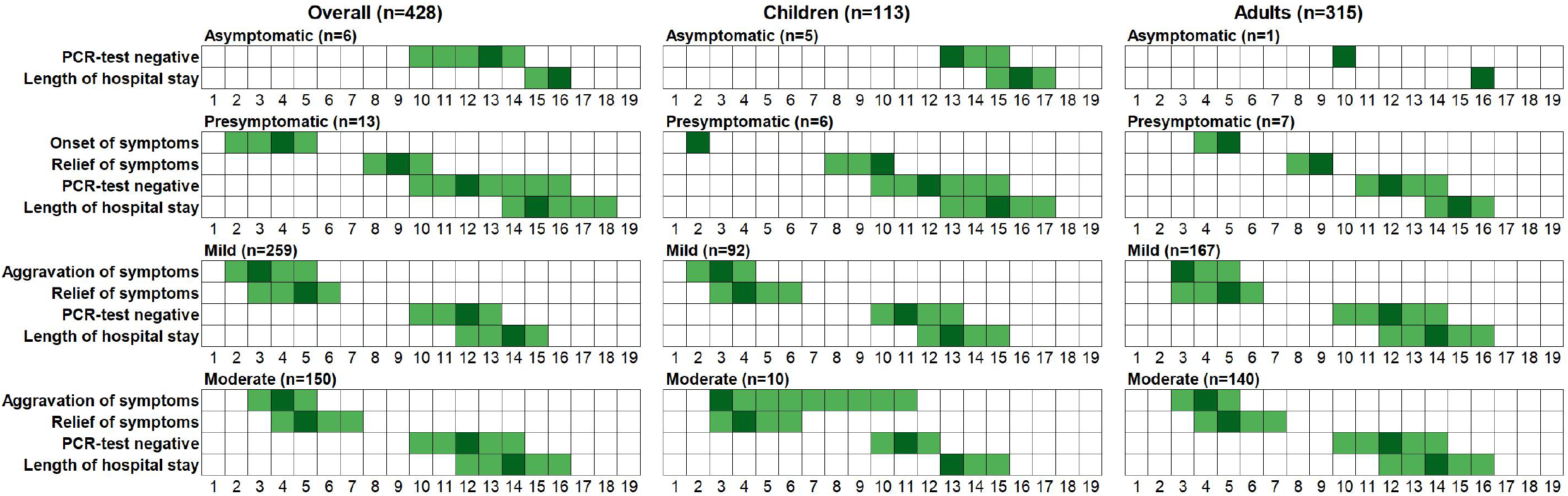
Course intervals of the SARS-CoV-2 Omicron infection in asymptomatic, presymptomatic, mild and moderate subject, stratified by age. The numbers in x-axis are days. The grids are filled with median and interquartile range. Dark color denotes medians and light color denotes the IQRs. Patient with severe disease (n=1) was excluded.

## Discussion

To our knowledge, this study reports the first population-based cohort of Omicron outbreak in China. CT-confirmed moderate Covid-19 had an overall prevalence of 35.0% on admission and 42.2% during hospital stay. The prevalence of moderate COVID-19 on admission and during hospital stay in children was 8.8% and 10.6%, whilst in adults 44.3% and 53.5%, respectively. The median length of hospital stay was 14.0 (IQR: 12.0, 15.0) days for all patients and the median interval of reversing positive PCR-test into negative was 12.0 (10.0, 13.0) days.

### Comparisons with other study findings

The situation in China and more specifically in Tianjin is different from the settings of many, maybe all settings of other papers in several aspects: Null COVID strategy in China, Chinese vaccine, whole population screening, strict quarantine, and strict nonpharmaceutical interventions (NPIs). Apart from keeping social distance, wearing masks, reducing public gatherings, additional NPIs in China included verification of health codes and travel record card (which were embedded on mobile phone apps) at all entrances of public buildings, and transformation from offline to online activities (teaching, meeting, etc.). This makes a direct comparison with other study findings difficult. Moreover, these study findings cannot compare with any other outbreak study with Omicron SARS-CoV-2 in China, because of the uniqueness of the outbreak in Tianjin. Therefore, the following comparisons with other study results are limited and needs a cautious interpretation.

### Study setting and course findings

The prevention and control of Covid-19 in China has been conducted on an ongoing basis since April 29, 2020, after the last hospitalized Covid-19 patient in Wuhan was discharged [13]. Afterwards, China has strictly adopted dynamic and targeted approaches all over the country to ensure early detection, quick response, targeted prevention and control, and effective treatment against Covid-19 [13], known as null-Covid strategy. This strategy ensured the fast tracing of all individuals who were infected by the Omicron variant and their close contacts, followed with strict quarantine and regular monitor to reduce further transmission. And this provided the possibility to calculate the course intervals using positive PCR-test, mostly the same day with hospital admission, as a beginning, which was unable to achieve in most previous studies. We found that the time intervals of reversing positive PCR-test into negative and the lengths of hospital stay were similar among different illness severity groups, which was probably a result of comprehensive interventions and treatments during hospital stay. These results need to be understood and interpreted cautiously. Based on our results, the time needed for reversing positive PCR-test into negative might be 2 weeks or longer in patients with Omicron infection, assuming the interventions or treatments were effective.

### Illness severity findings

Our results show a high prevalence of moderate Covid-19, while previous studies have focused more on the hospital admission rate, proportion of severe, critical and fatal diseases and demonstrated that Omicron patients were more likely to be mild, less hospitalized, less severe disease, and of lower mortality in South Africa [2,3], these findings were confirmed with data from Canada [4] and England [5]. In our study, the Omicron infection was most likely to be mild (over 50%) with no fatal cases observed, however, the proportion of moderate Covid-19 should not be neglected. It is plausible that the possibility of developing long-term sequelae of COVID-19 would be higher in moderate than in mild patients, which is in particular important for children. The intrinsic virulence of omicron may be not as mild as observed.

### Protective effect of vaccination

Covid-19 vaccines, in particular with a booster dose, has been reported associated with less infections [14], less severe illness [15,16], lower likelihood of being symptomatic [17], but the protection effect may wane with time [14] and be weakened via immune escape [18,19] and confounded by different levels of herd immunity. Notably, COVID-19 vaccines for children are currently unavailable or not yet offered in most countries [20], which leaves children more vulnerable to Omicron. In addition, previous infection of Covid-19 may also help build acquired immunity [21]. In the present study, nearly 80% of the study population received two or three doses of Chinese inactivated vaccines, and none of the patients had prior Covid-19 infection. The inactivated vaccines are different from mRNA vaccines in nature, which may result in different effectiveness against the Omicron variant, as none of them were initially designed for this variant, thus direct comparisons between our study and others were inappropriate. Although preexisting acquired or natural immunity normally plays an important role [22, 23], it is noteworthy to mention that the patients in our study were monitored in hospital since they were tested positive, preventive medicine and treatments according to symptoms were available for all patients, which might have also played an important role in reducing illness severity.

### Strengths and limitations

This study has several strengths, which are summarized as followed: This is an almost complete whole population study of the entire inhabitants of Tianjin, which allows a quantitative evaluation of disease courses and disease progression of SARS-CoV-2 Omicron infection. The present study traced all patients of the outbreak and included detailed information on clinical course started early since positive nucleic acid tests. Moreover, data on children have been insufficiently described in previous studies. A third strength owes to the use of CT scans to confirm the diagnosis of moderate disease.

This study also has several limitations. First, due to the unique characteristics of the setting of this study, the results might be presumably similar in other regions of China, but not in other parts of the world. Thus, this study has a limited generalizability, because the study population received inactivated vaccines, where mRNA vaccines were used in most of the settings of other publications. Second, adjustment for potential confounders were not applicable due to small numbers or low prevalence in children, although stratification analyses were conducted. Third, following the regulations of null-Covid strategy in China, all patients in the present study were hospitalized as opposed to most other populations in the world, the results may not correlate as well to other populations. However, this setting strengthens our confidence of describing real-world distribution of disease severity, as recruiting all positive patients is infeasible in most cases.

In conclusion, symptomatic Covid-19 was predominant in the Omicron outbreak in Tianjin, with high prevalence of moderate Covid-19 in adults and children. The course from positive PCR-test to negative took approximately 2 weeks. The findings suggest that moderate course of Covid-19 had unexpectedly higher proportion in the entire patient population, which is important in light of the current epidemic in China.

## Data Availability

All data produced in the present work are contained in the manuscript

## Statements and declarations

### Author Contributions

Zhengcun Pei had full access to all the data in the study and takes responsibility for the integrity of the data and the accuracy of the data analysis.

Yi Ren and Lixia Shi contributed equally to this article.

Concept and design: Zhengcun Pei, Yi Ren, Lixia Shi, Qi Wu

Acquisition, analysis, or interpretation of data: All authors.

Drafting of the manuscript: Zhengcun Pei, Yi Ren, Lixia Shi

Critical revision of the manuscript for important intellectual content: All authors

Statistical analysis: Zhengcun Pei

Data collection: Yi Xie, Chao Wang, Wenxin Zhang, Feifei Wang, Haibai Sun, Lijun Huang, Yuanrong Wu, Zhiheng Xing, Wenjuan Ren

### Ethics Approval

This study was performed in line with the principles of the Declaration of Helsinki. Approval was granted by the institutional review board at Haihe Hospital (Approval number: 2022HHWZ-002), the written informed consent was waived owing to the use of retrospective data.

### Data Availability

The data underlying this article cannot be shared publicly for the privacy of individuals that participated in the study. The data will be shared on reasonable request to the corresponding author.

